# Cancer and Circulatory Disease Risks for the Largest Solar Particle Events in the Space Age

**DOI:** 10.1101/2023.08.14.23294050

**Authors:** Francis A. Cucinotta, Sungmin Pak

## Abstract

In this paper we use the NASA Space Cancer Risk (NSCR version 2022) model to predict cancer and circulatory disease risks using energy spectra representing the largest SPE’s observed in the space age. Because tissue dose-rates behind shielding for large SPE’s lead to low dose-rates (<0.2 Gy/h) we consider the integrated risk for several historical periods of high solar activity, including July-November, 1960 events and August-October 1989 events along with the February 1956 and August 1972 events. The galactic cosmic ray (GCR) contribution to risks is considered in predictions. Results for these largest historical events show risk of exposure induced death (REID) are mitigated to < 1.2% with a 95% confidence interval with passive radiation shielding of 20 g/cm^2^ aluminum, while larger amounts would support the application of the ALARA principle. Annual GCR risks are predicted to surpass the risks from large SPEs by ∼30 g/cm^2^ of aluminum shielding.

## Introduction

Solar particle events (SPE) consisting largely of protons, with energies from below 10 MeV to several hundred MeV, present risks for both acute and late radiation health effects. Detailed measurements of SPE have been made since solar cycle number 19 (SSN-19) (1954-1964) [1-3] including thru the recent SSN-24 (2008-2019) [3]. The total fluence, fluence-rate, time profile, and energy spectra are known to be highly variable for each event. Large SPE’s considered herein are defined as events with integral proton fluences above 30 MeV, F(>30 MeV) > 10^8^ per cm^2^. We note that particles with energies below 30 MeV/u have insufficient range to be of concern for critical organ exposures behind minimal shielding. The frequency of large events displays significant variability in specific solar cycles with a larger number occurring in SSN-19 and SSN-22 compared to SSN’s 20, 21 and 24. In addition the temporal profile of solar events is highly variable with rapid onset (<4 h) to high proton intensity for the August of 1972 events and slower onset (>8 h) for the October of 1989 event.

The above-mentioned aspects of variability in the characteristics of SPE, including time of event occurrence, temporal profile, and total fluence and energy spectra, leads to issues on possible mitigation approaches. In previous work it was shown that acute radiation health risks from SPE are extremely unlikely [4-6] to occur for radiation shielding >5 g/cm^2^ (aluminum) because of the reduced doses and dose-rate. Thus, acute radiation risks are largely an issue for extra-vehicular activities (EVA). The risks of late effects are a major concern for SPE risks leading to a focus on the ability of spacecraft shielding to mitigate health risks. However, in this report we show that the use of typical amounts of passive radiation shielding in spacecraft (∼20 g/cm^2^) reduces cancer risks to acceptable levels for all major periods of solar activity of SSN 19-24.

The purpose of this report is to document the risk of late effects of cancer and circulatory diseases using the NASA Space Cancer Model (version NSCR-2022) [7-10]. For model proton spectra we use the analysis of Tylka et al. [11,12] that performed a detailed analysis of large SPE with ground-level enhancements on (GLE) on Earth to estimate the high-energy portion of the spectra and validated the method by comparing to satellite data at lower energies (≤200 MeV). Their analysis made improved estimates of Earth magnetic field rigidity cutoffs and data for a large number of neutron-monitors at various geographical locations to estimate proton energy distributions.

Uncertainty analysis in radiation risk predictions are of importance because of the sparsity of epidemiology and radiobiology data, especially for high linear energy transfer (LET) radiation [1,7]. A risk prediction is not scientifically valid without an assessment of the uncertainty in the prediction [1,7]. The use of an effective dose maybe acceptable for low risk levels well below an acceptable risk limit, however as a limit is approached its use is confounded by large uncertainties thus requiring their evaluation. The NASA Space Cancer Risk (NSCR) model has evolved for nearly 25 years to consider changes in epidemiology data and new results from space radiobiology research and performs a detailed uncertainty analysis using Monte-Carlo sampling over probability distribution functions (PDFs) that represent possible values of factors that enter into risk assessment calculations. In recent work the effects of sex, age, and racial or ethnic group are considered in risk predictions [9,10]. We also consider never-smokers (NS) as more representative of astronauts than an average population made-up of current, former and never smokers. The cancer risks for a population of white females were shown to be the highest risk group. Therefore, we consider SPE risk scenarios of 35-year old NS female astronauts with and without an annual galactic cosmic ray (GCR) contribution.

## Methods

We briefly summarize the method developed to predict the risk of exposure induced cancer (REIC) and risk of exposure induced death (REID) for space missions and associated uncertainty distributions [7-10]. The instantaneous cancer incidence or mortality rates, λ_I_ and λ_M_, respectively, are modeled as functions of the tissue averaged absorbed dose *D*_*T*_, or dose-rate *D*_*Tr*_, sex, age at exposure *a*_*E*_, and attained age *a* or latency *L*, which is the time after exposure *L=a-a*_*E*_. The λ_I_ (or λ_M_) is a sum over rates for each tissue that contributes to cancer risk, λ_IT_ (or λ_MT_). These dependencies vary for each cancer type that could be increased by radiation exposure. The rates are then combined with a function defining radiation quality and dose-rate effects to predict REID or REIC and described previously [7-10]. Epidemiology data for background cancer, circulatory disease and life-table rates, and low LET epidemiology data are described in previous reports [9,10].

NSCR-2022 used a parametric function denoted as *R*_*QF*_ that describes the radiation quality and dose-rate dependence of hazard rates (λ_I_ or λ_M_) for particles of a given energy and charge number. The *R*_*QF*_ is estimated from relative biological effectiveness factors (RBE’s) and dose-rate modifiers determined from low dose and dose-rate particle data relative to acute γ-ray exposures for doses of about 0.5–3 Gy [7]. The quality function, *R*_*QF*_, considers several contributions. The first representing an ion track’s penumbra of energetic δ-rays (electrons), which are assumed to follow a linear dose response estimated from γ-ray epidemiology studies. The dose-rate modifier is assumed to be influenced by results from experimental models used in RBE determinations, and similar to estimates of a dose and dose-rate reduction effectiveness factor (DDREF)). This factor adjusts the penumbra like term. The second term represents an ion’s track core of ultra-high ionizations whereby no dose-rate modifier is assumed. Radiobiological experiments that vary the charge number and velocity of an ion were considered to determine the relative contributions of the penumbra and core components. In addition, a 3^rd^ term in *R*_*QF*_ represents the contribution of non-targeted effects (NTE) is considered for high LET ions [8].

In this approach the scaling factor for the penumbra-like and core-like terms of low and high ionization densities, respectively is based on the following parametric function denoted as targeted effects (TE) terms:

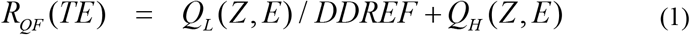

where

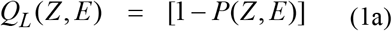

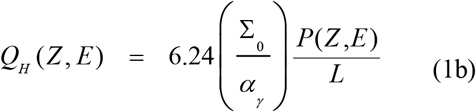

with the parametric function [7]:

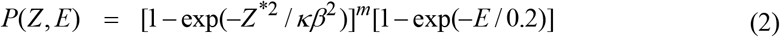

where *E* is the particles kinetic energy per nucleon, *L* is the LET in keV/μm, *Z* is the particles charge number, *Z\** is the effective charge number, and *β* is the particles speed relative to the speed of light. An ancillary condition is used to correlate the values of the parameter κ as a function of *m* as:

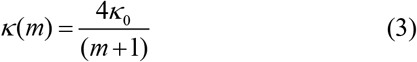

where κ_0_ is the value for the most likely value *m*=3. This constraint fixes the peak effectiveness with kinetic energy for each heavy ion charge group in the model to be consistent with results from experiments. The three model parameters (Σ_0_/α_γ_, κ and *m*) in Eq. (1-3) are fit to radiobiology data for tumors in mice or surrogate cancer endpoints as described previously [7,13] as shown in **Table 1**. Distinct parameters are used for estimating solid cancer and leukemia risks based on estimates of smaller RBEs for acute myeloid leukemia and thymic lymphoma in mice compared to those found for solid cancers [7].

**Table 1.** Space Radiation Quality Factor Model Parameters with standard deviation (SD) in parameter estimate.

| Model<br>Parameter | High Z ($Z>4$) | Low Z ($Z\leq 4$) |
| --- | --- | --- |
| Slope<br>parameter, m | $3\pm 0.5$ | $3\pm 0.5$ |
| $\kappa$ | $624\pm 69$ | $1000\pm 150$ |
| **Solid Cancer** |  |  |
| $\Sigma_0/\alpha_\gamma, \mu\text{m}^2\text{Gy}^{-1}$ | $(2897\pm 357)/6.24$ | $(2897\pm 250)/6.24$ |
| **Leukemia** |  |  |
| $\Sigma_0/\alpha_\gamma, \mu\text{m}^2\text{Gy}^{-1}$ | $(1750\pm 250)/6.24$ | $(1750\pm 250)/6.24$ |
| **Parameters for NTE model** |  |  |
| $\eta_0/\alpha_\gamma, \text{Gy}^{-1}$ | $(6\pm 3)\times 10^{-5}$ | $(6\pm 3)\times 10^{-5}$ |
| $\eta_1$ | $833\pm 200$ | $1000\pm 250$ |
| $A_{\text{bys}}, \mu\text{m}^2$ | $1000\pm 333$ | $1000\pm 333$ |

### Transport Codes, SPE and GCR Environments

Calculations are made using the BRYNTRN code for SPE [14,15] and HZETRN code [16,17] for GCR. Nuclear fragmentation including the production of target fragments in tissue are considered for SPEs and GCR. For radiation transport in spacecraft materials and tissue estimates of the particle energy spectra, *ϕ*_*j*_*(E)* for 190 isotopes of the elements from Z=1 to 28, neutrons, and contributions from pions are made as described previously [18]. We performed calculations for simple spherical geometries made-up of aluminum shielding of increasing thickness. Organ shielding is represented by water equivalent material using the CAM and CAF combinatorial geometry model for males and females, respectively [19,20]. For heart exposure estimates we use the MAX/FAX model shielding distribution based on CT scan data [21].

Integral distributions in proton rigidity, *R*, for a larger number of ground level enhancements (GLE) recorded by various neutron monitors were fit by Tylka et al. [11] with the so-called Band function in terms of particle rigidity:

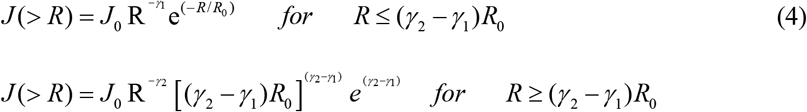

The Band functions parameters (*J*_*0*_, *R*_*0*_, *γ*_*1*_ and *γ*_*2*_) for the events considered are provided in **Table 2**. The function of Eq. (4) is transformed into a differential energy function of calculations described below.

**Table 2.** Band function parameters based on fits to data on energy spectra of several historically large SPEs from Tylka et al. [11,12].

| **Group #** | **Event** | **$J_0$, protons/cm<sup>2</sup>** | **$\gamma_1$** | **$\gamma_2$** | **$R_0$, GV** |
| --- | --- | --- | --- | --- | --- |
| **1** | 2/23/1956 | $8.79 \times 10^8$ | 0.584 | 5.04 | 0.3207 |
| **2** | 9/3/1960 | $1.24 \times 10^8$ | 0.319 | 5.56 | 0.1410 |
| **2** | 11/12/1960 | $7.54 \times 10^8$ | 1.154 | 6.54 | 0.2160 |
| **2** | 11/15/1960 | $1.78 \times 10^8$ | 1.595 | 7.0 | 0.2628 |
| **2** | 11/20/1960 | $3.07 \times 10^7$ | 1.112 | 5.21 | 0.1970 |
| **3** | 8/4/1972 | $1.45 \times 10^{15}$ | -3.636 | 7.95 | 0.0345 |
| **3** | 8/7/1972 | $6.34 \times 10^6$ | 3.26 | 6.27 | 0.298 |
| **4** | 8/16/1989 | $7.27 \times 10^7$ | 1.914 | 5.86 | 0.1845 |
| **4** | 9/29/1989 | $2.027 \times 10^{10}$ | -0.109 | 4.58 | 0.0945 |
| **4** | 10/19/1989 (GLE) | $1.22 \times 10^9$ | 0.528 | 5.81 | 0.1621 |
| **4** | 10/19/1989 | $9.09 \times 10^9$ | 0.911 | 4.43 | 0.08435 |
| **4** | 10/22/1989 | $1.09 \times 10^9$ | 1.226 | 7.25 | 0.1352 |
| **4** | 10/24/1989 | $4.42 \times 10^7$ | 2.176 | 5.565 | 0.385 |

As described previously [22] we modelled recent and historical data GCR spectral data using the approach suggested by Wiedenbach et al. [23] where solar modulation is described by the forced-field approximation [24] fit to CRIS data [25] and historical balloon measurements (see ref [22]). The model uses the Voyager I spectral data [26], which fixes values of the local interstellar spectra (LIS). The values at lower energies (up to a few hundred MeV/u) are an important factor in data fits because of the overlap with solar modulation in this energy region. For a particle of momentum, p (MeV/c) and kinetic energy per mass unit, E (MeV/u), the LIS for a given element of charge number, Z and mass number, A is modeled with the often-used high energy form with a low energy correction

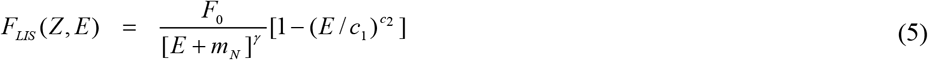

where *m*_*N*_ is the nucleon mass. The near-Earth spectra for modulation value, Φ, under the forced-field approximation is given by

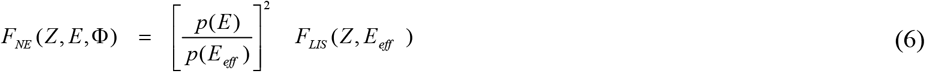

where the E_eff_ is an effective energy that approximately accounts for the energy shift due to particle transport in the heliosphere as determined by the solar modulation parameter, Φ(MV):

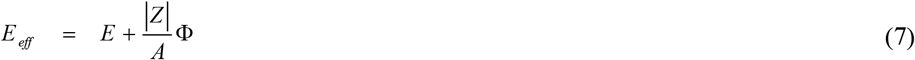

A χ^2^ minimization taken into account the measurement uncertainty is made to estimate the model parameters in Eq. (5) for each charge group using the Voyager I, CRIS data, and balloon and satellite data. For each primary element isotopic ratios are used to model the isotopic dependence of the primary GCR [27].

## Results

**Figure 1** shows the energy distributions for several historical large SPEs that are considered. Because the duration of exposures for individual events is from >8 h to several days we assumed organ doses behind shielding are of low dose-rates (<0.2 Gy/h) and grouped series of events over a 3-month period for risk predictions. We considered predictions for the Feb 23, 1956 event, the series of the 4 events from September to November 1960, the August 4-7, 1972 event, and the series of 5 events from August thru October of 1989 (note **Table 1** uses fits to the GLE portion of the October 19, 1989 event separately from the later portion of this event). Tylka et al. [11] note that their fits of the 1972 event underestimate the fluence at 10 MeV or lower energies, however this energy region makes a negligible contribution to organ exposures. The 1989 series is largest in the important intermediate energy range of 100 to 300 MeV protons [6], however is surpassed at higher energies by the February 23, 1956 event.

**Figure 1.**
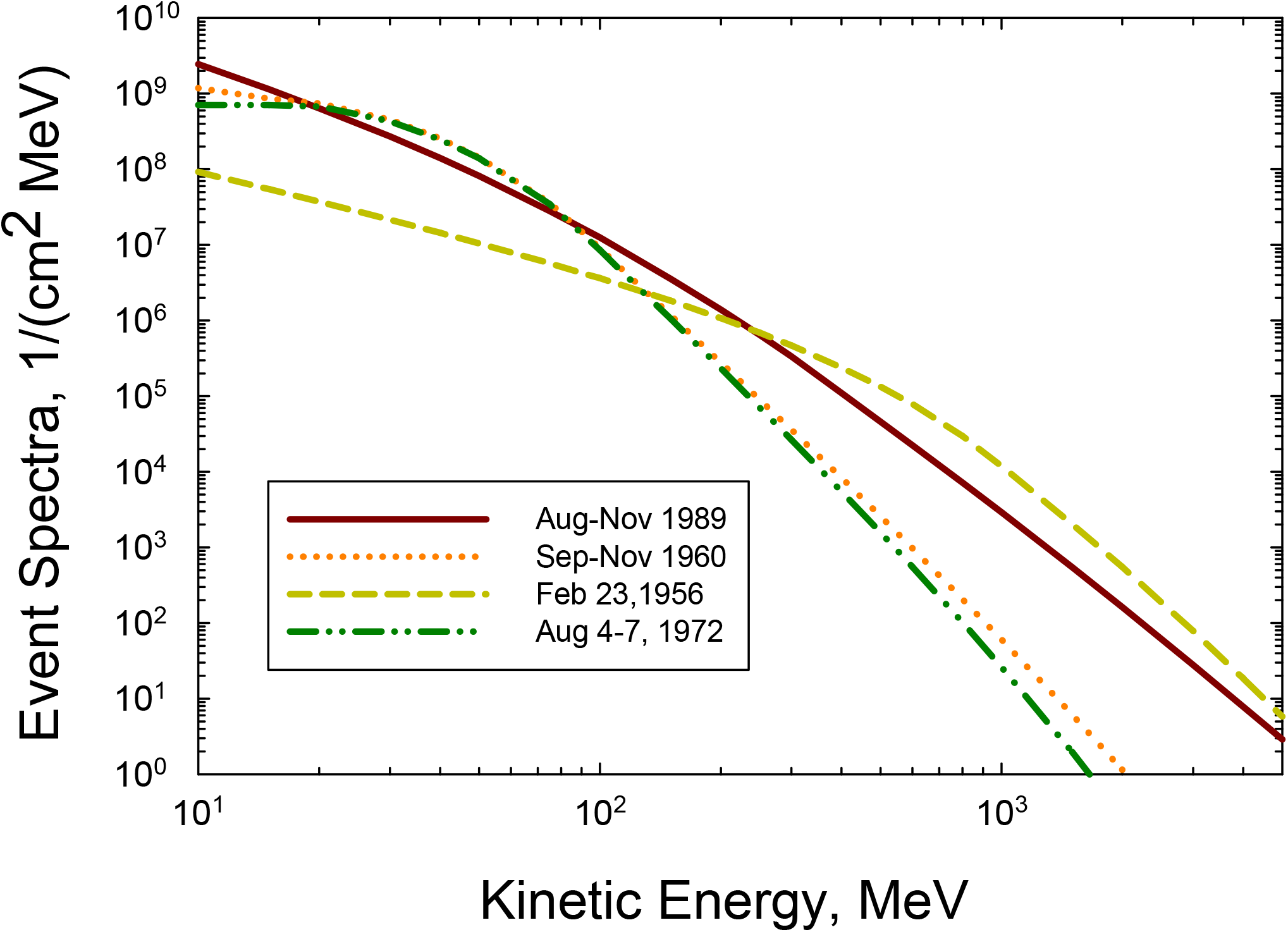
Solar proton energy spectra for several major SPE or series of SPEs using spectra fits from Tylka et al. [11,12].

For the GCR environment, noting that large SPEs have occurred a few years prior or after solar maximum [1], we performed calculations with a solar modulation parameter of 800 MV. For GCR predictions, NSCR has used tissue specific organ doses and a tissue average fluence spectra as a function of the track structure descriptor, Z^*2^/β^2^. **Figure 2** shows spectra for several organs in females that verifies the accuracy of the use of average tissue spectra for GCR. The GCR spectra decrease rapidly at large Z^*2^/β^2^, however we note that the risk per particle is expected to increase as ∼ (Z^*2^/β^2^)^3^ as indicated by Eq. (2). In contrast to GCR, SPEs have a rapid decrease in fluence and dose with increasing shielding due to the large contribution of low to medium energy protons. Therefore, a large variation in spectra for different tissues occurs. **Figure 3** shows these variations for the 1956 event and the 1960 series of events. **Figure 3** indicates a decreasing fluence with increasing depth along with differences in the spectral shapes between tissues. The present NSCR-2022 uses tissue specific spectra for risk predictions from both GCR and SPE.

**Figure 2.**
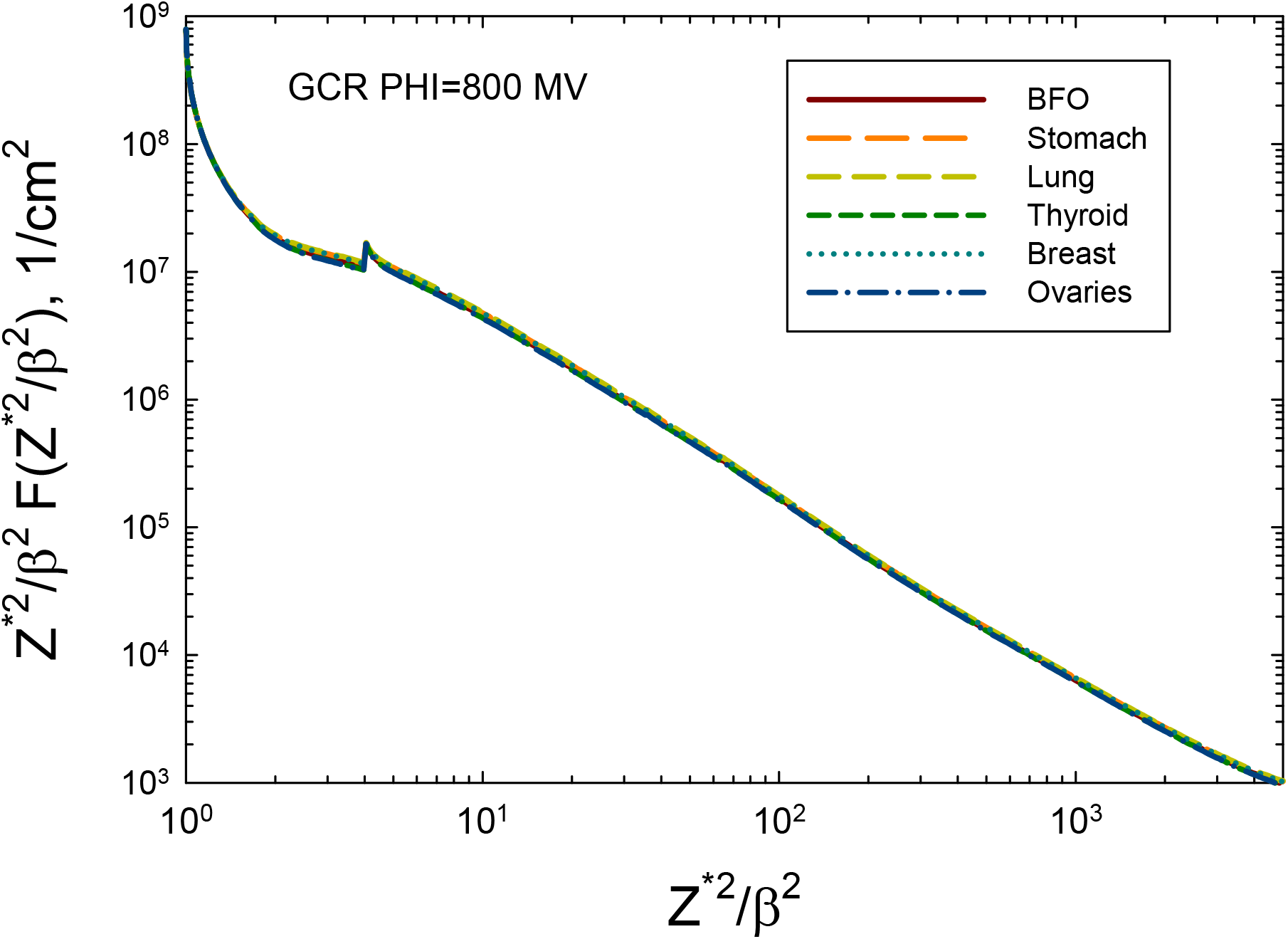
GCR fluence spectra behind 20 g/cm^2^ aluminum shield at several tissues in females, including the average at several blood forming organ (BFO) sites. Calculations with the HZETRN code are for a modulation parameter of Φ=800 MV. Results are shown as (Z^*2^/β^2^) x F(Z^*2^/β^2^) versus Z^*2^/β^2^.

**Figure 3.**
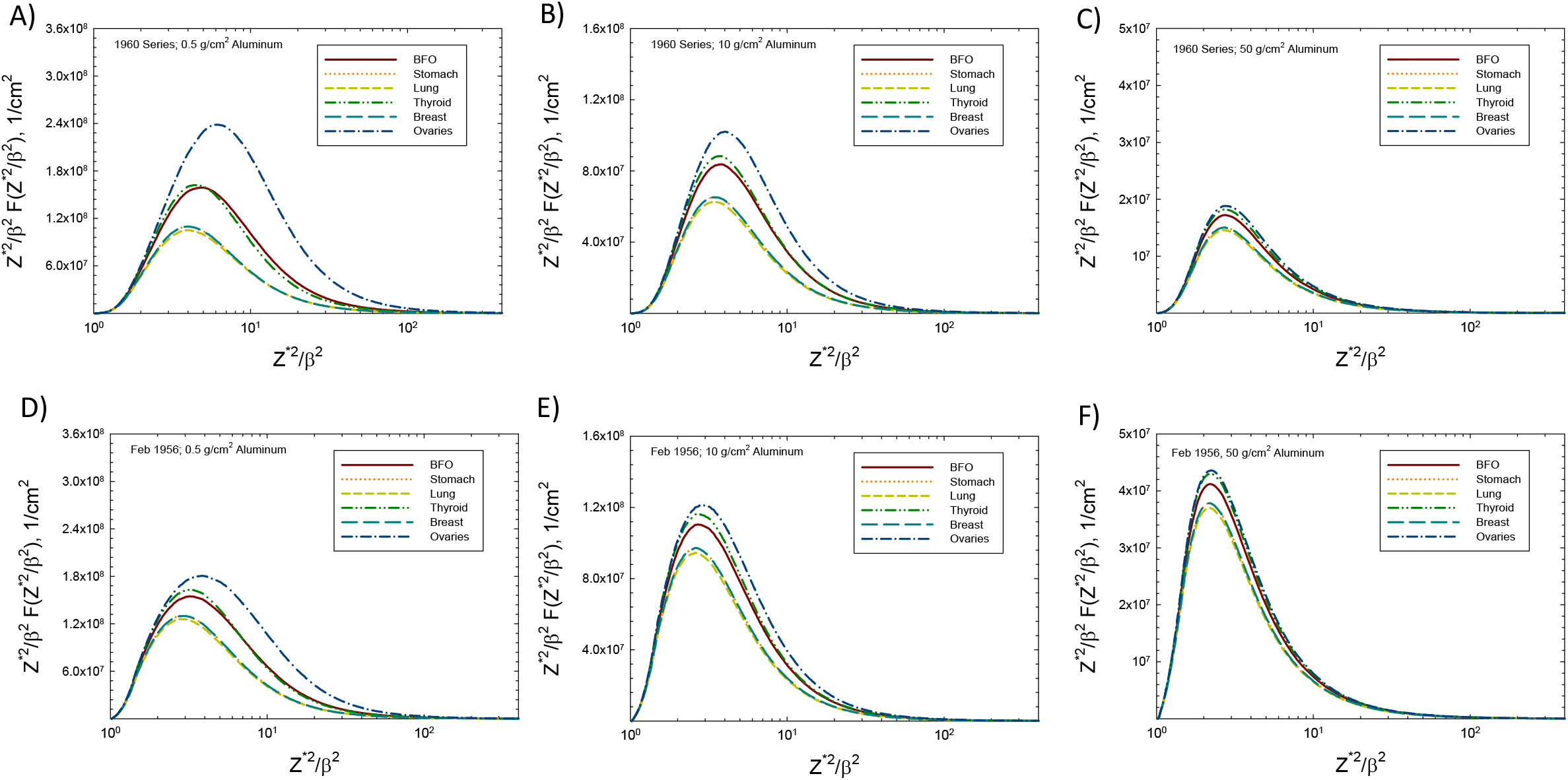
BRYTRN code predictions of tissue specific lethargy spectra of track structure parameter Z^*2^/β^2^ for the 1956 event and 1960 series of events. Panels A, B, and C are for the 1960 series for aluminum shields of 0.5, 10 and 50 g/cm^2^. Panels D, E, and F are for the February 23, 1956 event for aluminum shields of 0.5, 10 and 50 g/cm^2^.

In **Figure 4** we show depth-dose curves in aluminum for the 1989 series of SPEs for the colon and breast, which are chosen to illustrate possible variations between tissues important for cancer risk. Absorbed doses and dose equivalents, *D*_*T*_ for solid cancer risk, *H*_*T*_(Solid), respectively are shown. Values for *H*_*T*_ (Solid) are ∼40% and ∼20% higher for breast compared to colon for 5 g/cm^2^ and 50 g/cm^2^, respectively. For GCR the values for breast and colon differ by <5% (result not shown), and **Figure 4** shows results for annual GCR breast *D*_*T*_ and *H*_*T*_*(Solid)*. Due to secondary particle production GCR doses increase slowly with shielding for a solar modulation of 800 MV. At solar maximum a larger increase would occur, while near solar minimum some reduction is expected especially at smaller shielding depths because of the increased contribution from lower energy primary GCR ions. These results show annual GCR become dominant over major SPE organ dose equivalent at ∼20 g/cm^2^. However, some differences in these results would occur for lower or higher solar modulation parameter assumptions along with specific mission length considerations. Results for *H*_*T*_*(leukemia)* using average blood forming organ (BFO) or non-cancer tissue dose, *G*_*T*_ using heart and brain shielding and RBE’s recommended by the NCRP [28,29] are intermediate between the values of *D*_*T*_ and *H*_*T*_*(solid)*.

**Figure 4.**
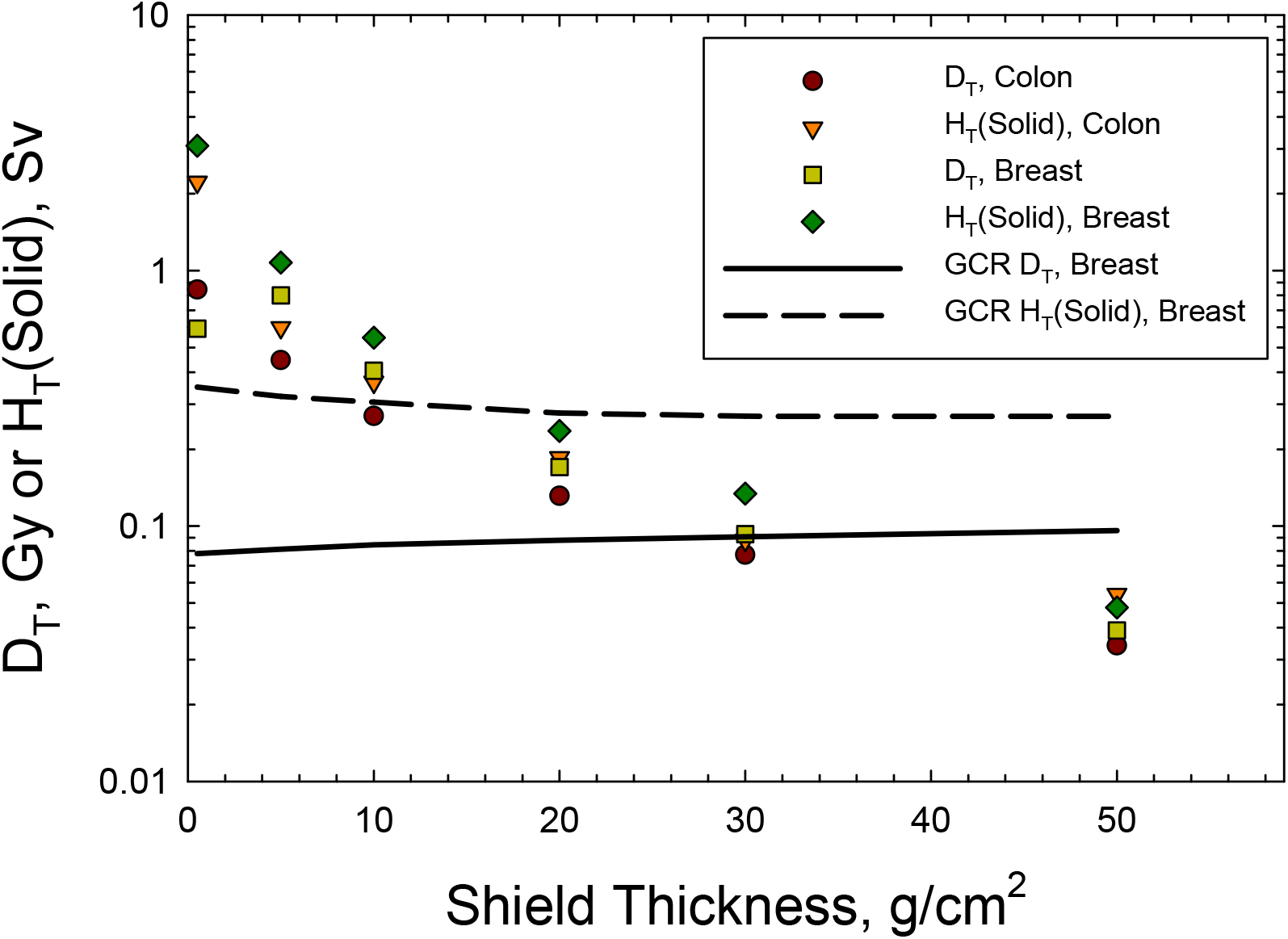
Predictions of colon and breast absorbed doses, D_T_ and solid cancer dose equivalent, H_T_(Solid) for combined August to October, 1989 events and GCR annual breast D_T_ or H_T_(Solid). For GCR values for colon and breast agree to within 5% and therefore results for colon are not shown.

**Table 3** shows predictions for the SPE’s considered of REIC, and REID for cancer and circulatory diseases for shielding of 5, 20 and 50 g/cm^2^ of aluminum. Upper 95% confidence intervals are about 2-fold higher than the mean estimate for all cases. The Monte-Carlo propagation of uncertainties made by NSCR-2022 includes models of PDFs for epidemiology data, transport code particle spectra predictions, radiation quality and dose-rate modifiers. However, uncertainties in the estimates of the SPE energy spectra have not been considered. Results show that 20 g/cm^2^ of aluminum shielding reduces REID values below 1% with upper 95% confidence intervals to < 1.2%. Circulatory risks are much smaller than cancer risks for age 35-y females, however their fractional contribution increases with age and for males [9,10]. In **Table 4** we show results for annual GCR risks for a solar modulation parameter of 800 MV, and risks for GCR combined with the 1956 event and 1989 series of events. For large SPEs or series of large events, annual GCR exposures provide an equivalent risk by ∼30 g/cm^2^ of aluminum shielding and dominate risks at larger shielding values.

**Table 3.**
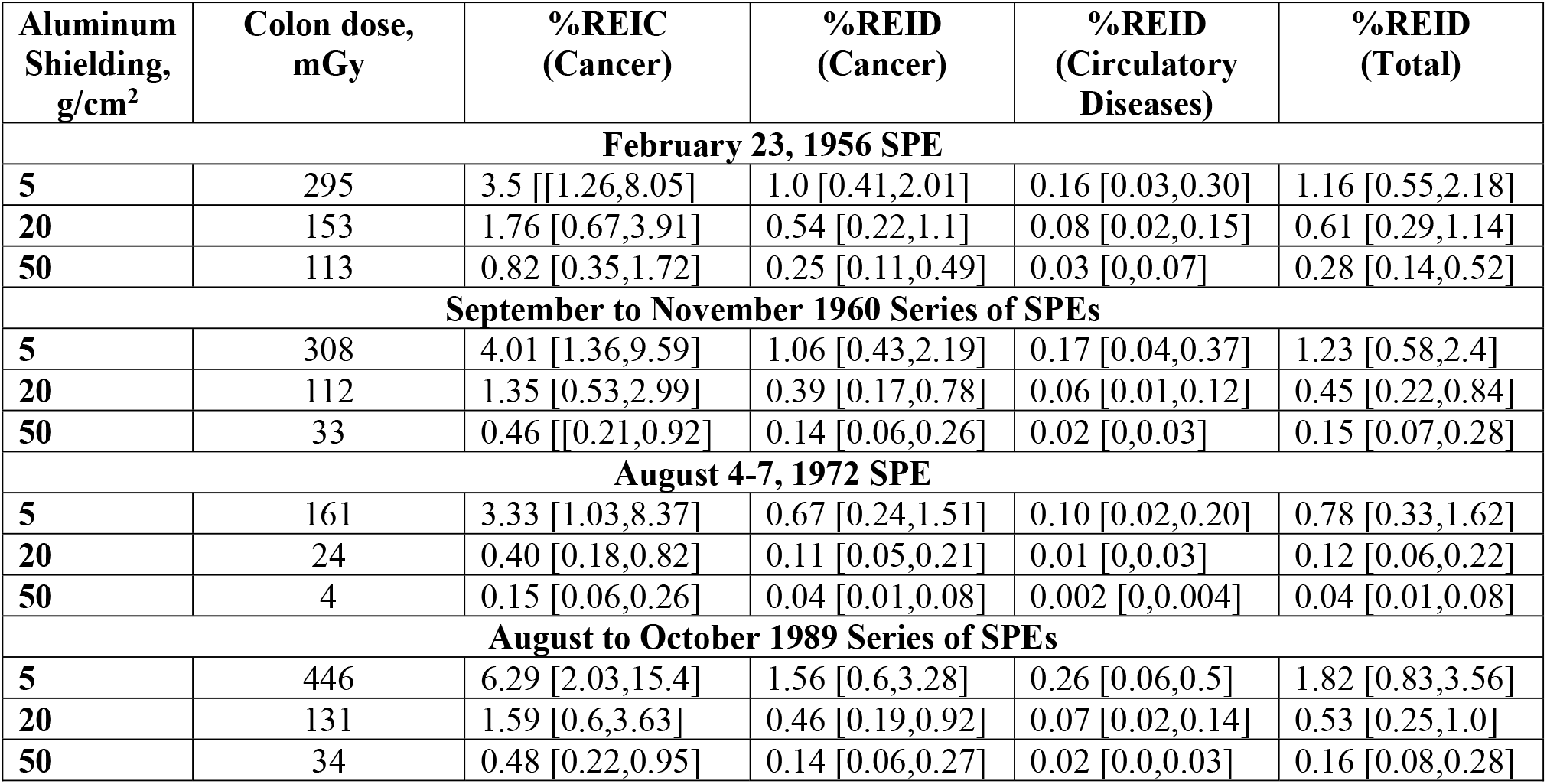
Predictions of cancer and circulatory disease risk and 95% confidence intervals for annual GCR for several historically large SPEs. Predictions for age 35-y white female never-smokers are shown for several thickness of aluminum shielding. REIC is risk of exposure induced cancer and REID is risk of exposure induced death.

**Table 4.** Predictions of cancer and circulatory disease risk and 95% confidence intervals for annual GCR for solar modulation Φ=800 MV, and GCR combined with February 23, 1956 or August to October, 1989 series of SPE. Predictions for 35-y old white female never-smokers are shown for several thickness of aluminum shielding. REIC is risk of exposure induced cancer and REID is risk of exposure induced death.

| Aluminum Shielding, g/cm<sup>2</sup> | Colon dose, mGy | %REIC (Cancer) | %REID (Cancer) | %REID (Circulatory Diseases) | %REID (Total) |
| --- | --- | --- | --- | --- | --- |
| **Annual GCR** |  |  |  |  |  |
| **5** | 81 | 0.95 [0.40,2.0] | 0.30 [0.13,0.58] | 0.05 [0.01,0.09] | 0.34 [0.17,0.63] |
| **20** | 88 | 1.03 [0.43,2.18] | 0.32 [0.14,0.63] | 0.04 [0.01,0.08] | 0.37 [0.18,0.68] |
| **50** | 96 | 1.15 [0.48,2.44] | 0.36 [0.16,0.71] | 0.04 [0.01,0.08] | 0.41 [0.20,0.75] |
| **February 23, 1956 SPE and Annual GCR** |  |  |  |  |  |
| **5** | 376 | 4.34 [1.56,10.0] | 1.12 [0.53,2.58] | 0.20 [0.04,0.39] | 1.46 [0.71,2.78] |
| **20** | 241 | 2.68 [1.01,5.98] | 0.83 [0.35,1.64] | 0.12 [0.03,0.23] | 0.95 [0.46,1.76] |
| **50** | 209 | 1.86 [0.73,4.09] | 0.59 [0.25,1.16] | 0.08 [0.02,0.15] | 0.66 [0.32,1.24] |
| **August to October 1989 Series of SPEs and Annual GCR** |  |  |  |  |  |
| **5** | 527 | 7.18 [2.36,17.3] | 1.84 [0.73,3.86] | 0.31 [0.07,0.59] | 2.15 [0.99,4.18] |
| **20** | 219 | 2.51 [0.92,5.62] | 0.75 [0.32,1.49] | 0.12 [0.03,0.23] | 0.87 [0.42,1.61] |
| **50** | 130 | 1.42 [0.57,3.07] | 0.45 [0.19,0.88] | 0.06 [0.01,0.12] | 0.51 [0.25,0.94] |

## Discussion and Conclusions

In this report we considered population-based cancer and circulatory disease risk predictions for age 35-y white female never-smokers. This population is chosen for their higher cancer risks compared to males and other racial or ethnic groups based on previous work [9,10]. Radiation cancer risks are expected to decrease with age of exposure, while circulatory disease risks vary much slower with age and are larger for males compared to females. A main conclusion of this report is cancer and circulatory risks from major SPE’s or a series of events over a few months are effectively mitigated with average aluminum shielding found in most spacecraft. Shielding augmentation using polyethylene or water could also be provided in “storm shelter” type configurations to practice the principle of as low as reasonably achievable (ALARA). Our considerations of major SPEs then ensure that other events in the space age would also be mitigated. The energy distribution using the Band function fits [11,12] led to lower fluxes at high energy for the August 1972 event from King spectra [30] and the February 1956 event spectra from Foelshe [31], which were often used in past model calculations [1,14]. For the August 1972 event the Band function provides a poor fit <10 MeV, which is an important energy region for micro-electronics damage on satellites.

Non-targeted effects were not considered herein because they are expected to play a small role for SPEs that are dominated by low LET protons. NTEs are predicted to increase GCR solid cancer risks by about 2-fold compared to the predictions of **Table 4** for GCR [8,32]. Future work will consider NTE for SPEs and also take a closer look at neutron contributions to SPE organ doses and spectra using Monte-Carlo transport codes.

Risks on the lunar surface are reduced by 2-fold due to the moon’s solid body with a small albedo component due to solar particle interaction with the lunar regolith. Exposures near Mars would be reduced from observations near Earth with most models assuming an inverse-square radial gradient for particle fluence [1]. On the surface of Mars, protection is due to the solid body (2-fold reduction) and the Martian atmosphere which provides a vertical column of 18 g/cm^2^ of CO_2_ shielding. Other considerations include trajectories to Mars where a possibility of a Venus swing-by trajectory to Mars would reduce warning times and possibly increase particle fluxes. However, we expect our main conclusion for shielding in deep space to be valid for lunar or Mars surface operations.

The possibility of a larger event than observed in the space age has been discussed for many years [1] and continues to be analyzed [33]. Fluence spectra estimates are limited on information at higher energies (>200 MeV), which are needed for risk predictions as a function of shielding depth. Nevertheless, we expect an extremely rare event with several times the February 23, 1956 fluence (with same spectral shape) or larger fold increase for a softer event spectrum would be mitigated below a REID of 3% risk by passive radiation shielding locations that are found in most spacecraft. Exposures during EVA’s are larger than inside spacecraft [34]. Also, considerations of helium [35] or heavy ion [36] component of SPE’s are needed for EVA’s and light shielding albeit they provide small contributions at larger depths. Acute radiation risk then become possible, while the main mitigation for EVA’s continues to be real-time monitoring of the radiation environment, possibly enhanced by event forecasting, in order to alert crews to move to shelter.

## Data Availability

All data produced in the present study are available upon reasonable request to the authors

## Acknowledgements

No funding was received for this research.

